# Optimising the prevention, detection and management of diabetes distress in adults with type 1 diabetes: Feasibility study protocol (D-stress feasibility study)

**DOI:** 10.64898/2026.08.17.26360546

**Authors:** C Fabian-Therond, S Ahuja, I Papachristou Nadal, RIG Holt, S Watson, S Hussain, P Choudhary, RA Ajjan, R Harris, M Peck, J Mohammadi, S Sims, F Fiorentino, M Due-Christensen, J Huber, L Fisher, K Hardenberg, M Stadler, Huajie Jin, J Halliday, J Sturt, the D-stress study collaborators

**Author notes:** **Correspondence to:** Dr Clara Fabian-Therond.

## Abstract

**Introduction:** Diabetes distress describes the psychological and emotional burden of living with diabetes and is associated with reduced self-management and adverse diabetes outcomes. Clinical guidelines recommend routine assessment and management of diabetes distress, but this is not always implemented. Therefore, there is a need to develop approaches to deliver emotional health support in routine clinical care more effectively.

We describe here the protocol for a study to I) assess the feasibility of implementation of the D-stress Pathway, comprising Enhanced Usual Care (EUC) and an online, group-based, psychological diabetes distress reduction intervention called REDUCE, ii) evaluate the feasibility of the study protocol iii) detect an effect signal of diabetes distress score and Interstitial Glucose Time in Range and iv) refine initial programme theories of how both interventions (EUC and REDUCE) work, for whom, and under what circumstances.

**Methods:** This feasibility study includes a multicentre trial within a cohort design (TWICs) where sites have a staggered exposure to the interventions alongside a realist process evaluation. Four UK NHS diabetes services will recruit 80 adults with type 1 diabetes (≥1 year) using continuous glucose monitoring (CGM) (≥3 months). All participants will receive EUC and provide monthly data over 7 months on diabetes distress (measured by the Type 1 Diabetes Distress Assessment System (T1DDAS) and interstitial glucose measured by using continuous glucose monitoring. Participants with elevated diabetes distress, will be offered the six-week, group-based, online REDUCE intervention plus EUC, compared to EUC alone. Up to twenty participants with type 1 diabetes, ten family members/friends, sixteen healthcare professionals delivering EUC and five REDUCE facilitators will be interviewed to explore their experience of receiving training and delivering the D-stress Pathway. Up to 20 EUC consultations and REDUCE sessions will be observed.

**Analysis:** Feasibility will be assessed against pre-specified progression criteria and analysed descriptively using summary statistics. Primary outcomes include baseline level of diabetes distress, recruitment rate, intervention uptake, and data completeness, which will be analysed descriptively. Qualitative data will be analysed using framework analysis guided by realist programme theories developed for this study.

**Ethics:** Ethics approval has been granted by NHS Research Ethics Committee (REC) (Bromley REC: 25/LO/0469) and Health Research Authority obtained. All participants will provide informed consent.

**Trial registration no:** Registered at ClinicalTrials.gov number NCT07193446 on 26/11/2025.

**Protocol and statistical analysis plan:** The trial protocol and statistical analysis plan can be accessed at ClinicalTrials.gov.

**Strengths and Limitations:**

- Evaluates the delivery of an evidence-based care pathway for diabetes distress.
- Addresses recommendations from clinical guidelines.
- The study uses mixed methods approach to assess feasibility outcomes.
- Findings may have limited generalisability beyond the study settings and populations included.
- Estimates of the intervention impact may be limited.

**What is already known on this topic – summarise the state of scientific knowledge on this subject before you did your study and why this study needed to be done:** International clinical guidelines for the assessment and management of diabetes distress recommend comprehensive assessment with discussion as part of routine care with psychological interventions for managing elevated diabetes distress.

**What this study adds – summarise what we now know as a result of this study that we did not know before:** The study will assess the feasibility of delivering a care pathway for diabetes distress and its impact on emotional, behavioural and clinical health outcomes and identify the contexts and circumstances that support care pathway delivery and impact for people with type 1 diabetes. It will inform subsequent main study planning by ascertaining feasibility of recruitment, intervention delivery and determining recruitment and attrition rates.

**How this study might affect research, practice or policy – summarise the implications of this study**

The study will deliver an evidence-based care pathway, and a research protocol, for a subsequent main study evaluation. The main study will provide evidence for the care pathway which aligns with the 2026 EASD Clinical Guideline for the assessment and management of diabetes distress in routine care.

## INTRODUCTION

In the UK, 350,000 adults live with type 1 diabetes, with NHS costs of an estimated £1.2 billion per year largely associated with the treatment of diabetes-related complications (1–3). In addition to physical health complications, there is a high emotional burden associated with living with and self-managing type 1 diabetes. Therefore, there is a recognised need to address the detrimental impact type 1 diabetes has on the mental health and overall well-being of people living with the condition (4–6). Clinical guidelines for the management of type 1 diabetes include recommendations for routine psychological and emotional health assessments, and more recently the assessment and management of diabetes distress, to be embedded in care (7, 8). Diabetes distress refers to the psychological and emotional burden of living with diabetes and is associated with less engagement in self-management and adverse diabetes outcomes (9). Diabetes distress occurs when the level of a person’s self-care burden exceeds their capacity to cope with the demands of the clinical condition. Emotional regulation mechanisms have been shown to be a mediator in the development of distress (10).

Between 22-42% of adults living with type 1 diabetes report elevated levels of distress (11, 12). More recently, diabetes distress has been recognised as even more prevalent, with up to 80% of adults with diabetes reporting at least one clinically significant diabetes-related concern associated with emotional distress (13). The EASD clinical guideline states that healthcare professionals are encouraged to routinely ask about and assess diabetes distress during consultations (14). Good Practice Statements recommend discussing the emotional side of diabetes at every appointment, using open-ended questions, and employing valid, reliable assessment tools. Regular monitoring for diabetes distress is advised as part of the annual cycle of care, with results recorded in clinical notes and discussed openly with people with diabetes. When distress is identified, healthcare professionals should work together with the individual with diabetes to agree on next steps, ensuring follow-up support is tailored and person-centred and management interventions are explored and encouraged (14). Persistent unaddressed elevated diabetes distress can lead to diabetes burnout, characterised by states of exhaustion, detachment and powerlessness (15, 16). Common sources of diabetes distress for adults with type 1 diabetes are powerlessness, shame, health professional and family/social relationships, hypoglycaemia, management regime and food and eating distress (16). Advances in diabetes technology have heightened financial worries, healthcare quality, lack of diabetes resources and technology challenges to this list (17). For the management of elevated diabetes distress in adults with type 1 diabetes, the EASD guideline makes conditional recommendations to use psychological interventions, continuing glucose monitoring and automated insulin delivery systems to reduce diabetes distress (14, 18–20).

Despite clinical guidelines recommending emotional wellbeing in diabetes being incorporated into routine care (7, 21), coordinated care pathways for the detection, prevention and management of diabetes distress are underdeveloped in clinical practice and research.

Although international programmes have demonstrated that diabetes distress can be identified and managed through structured healthcare professional training (4, 22) and psychosocial interventions (23–25), these approaches have not been implemented as an integrated care pathway within routine NHS diabetes services. Consequently, despite national and international recommendations to assess and address diabetes distress, provision remains inconsistent and largely dependent on local services and clinician expertise.

This paper outlines the research protocol for the feasibility evaluation of the D-stress care pathway, which is part of the research programme being delivered between 2024-2029 entitled ‘Optimising the Delivery of Diabetes Distress Informed Care for Prevention, Detection and Management in Adults with Type 1 Diabetes: A Hybrid Effectiveness-Implementation Programme (D-stress Study)” co-funded by the UK NIHR and Diabetes UK (NIHR205441)(26).

### Development of the D-stress care pathway

Our research programme developed the D-stress care pathway (Fig 1), which is the focus of this feasibility study. The care pathway was developed to support NHS diabetes services to routinely identify and manage diabetes distress among adults with type 1 diabetes. It includes two interventions: (1) Enhanced Usual Care (EUC), and (2) REDUCE, both of which are described below (See ‘Interventions’). Development of the D-stress care pathway, EUC, and REDUCE, drew on internationally evaluated interventions supporting the routine comprehensive assessment of diabetes distress underpinned by diabetes health professional training (22, 27), prevention of diabetes distress and the management of elevated diabetes distress, from Australia, Denmark and the USA (28). Rapid realist reviews of literature were conducted to develop initial programme theories to explain how the two interventions in the D-stress pathway, Enhanced Usual Care and REDUCE, may work, for whom and in what circumstances (23, 29, 30).

**Fig 1.**
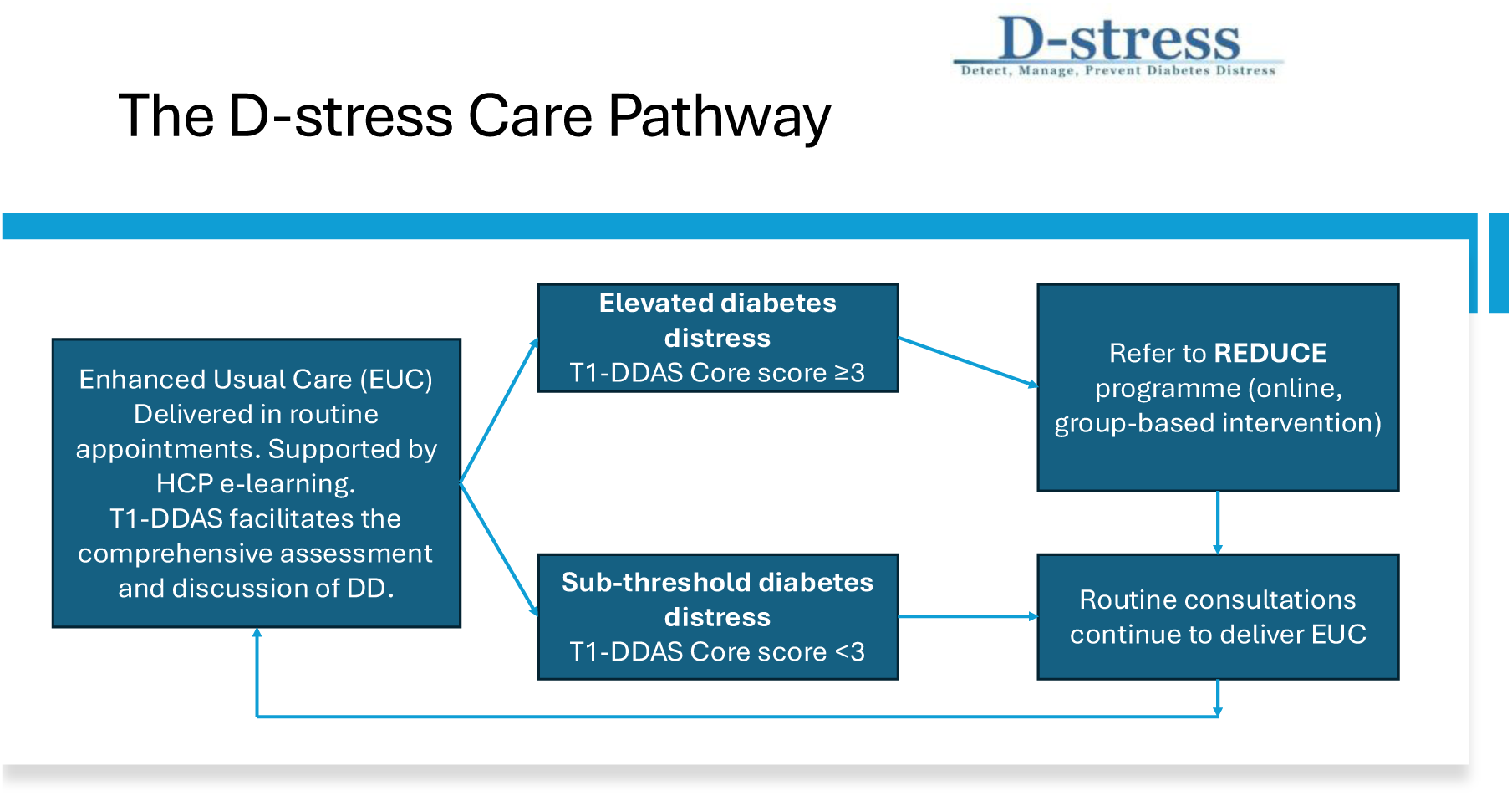
T1-DDAS Type 1 Diabetes Distress Assessment System and care pathway outline.

### Study aims

1. Assess the feasibility of delivery and implementation of the D-stress care pathway, comprising Enhanced Usual Care (EUC) and an online, group, psychological intervention called REDUCE.
2. Evaluate the feasibility of the study protocol.
3. Evaluate initial programme theories regarding how the intervention works, for whom, and under what circumstances.
4. Determine the feasibility of the research protocol for recruiting, randomising and retaining clinical sites and their patient population into a trial.
5. Determine the feasibility of data collection of key outcomes, including diabetes distress, comprehensive analysis of glucose levels using CGM, health-related quality of life and limited healthcare-use data, to inform the main trial and planned economic evaluation.

The findings of the feasibility study will inform protocol development for the ‘main trial’ evaluation study, to be conducted across 20 NHS diabetes centres during 2027 to 2028.

## METHODS AND ANALYSIS

### Study Design

The study is a 26-week, multicentre feasibility study. The design is a trial within a cohort (TWiC) cluster design (24), with staggered intervention exposure and an embedded mixed methods realist process evaluation (31, 32). The design will assess the feasibility of delivering the D-stress care pathway and collecting key outcome and implementation data to inform the design of the main trial, including the planned health economic evaluation. Realist process evaluation is based on realist philosophy, which assumes the “real-world” is filtered through human action, language, perception and culture that shape how clinical (and other types of social) interventions are implemented and embraced (or not) by users (33). By using this methodology, we aim to understand what aspects of the D-stress pathway work (and do not work), for whom, why and in what circumstances as it is being implemented and evaluated to inform the main trial, and to advance post-trial implementation. Evaluation of EUC for the prevention of elevated diabetes distress is the focus of aligned PhD research.

The TWiC design comprises a closed cohort of participants receiving Enhanced Usual Care. In the closed cohort design, participants are recruited at the beginning and remain until the end (34, 35). Participants with elevated diabetes distress (score ≥3) are offered entry into an embedded cluster trial, with staggered randomisation to the REDUCE intervention or continued Enhanced Usual Care. A staggered cluster RCT design is chosen for time efficiency, enabling intensive trial outcome data collection in a shorter time. This design minimises cluster exposure differences and ensures high ethical standards by providing swift access to the REDUCE intervention for those with elevated diabetes distress. A realist process evaluation runs alongside the cohort and the cluster trial and its mixed methods approach will evaluate the impact of both EUC and REDUCE for participants, routine care providers and REDUCE facilitators (33). The study design and protocol were informed by the SPIRIT (Standard Protocol Items: Recommendations for Interventional Trials) guidelines to ensure systematic and transparent specification of the trial design, procedures, and outcomes (36, 37).

### Research ethical and governance approvals

Ethics approval was granted by the London-Bromley Research Ethics Committee (study ID: 25/LO/0469) on the 17 July 2025. Amendments to the Protocol were made on 7^th^ April. 2026, 30^th^ September 2025, and 16^th^ September 2025. Protocol modifications were discussed with the study management and oversite committees and relevant parties across each site.

### Study monitoring

Protocol adherence, recruitment, consent, and data quality are monitored by the Chief Investigator and study management team. King’s College London and Guy’s and St Thomas’ NHS Foundation Trust, as co-sponsors, act as Data Controllers.

#### Programme Steering Committee (PSC)

Our study is supported by a PSC who meet at least annually and is made up of research and clinical experts in the field of Type 1 diabetes led by an Independent Chair and four independent members. It will consider and advise on the implications of any new evidence from both within the programme and other sources, or of any proposed changes to the programme.

#### Independent Data Monitoring Ethics Committee (DMEC)

The feasibility D-stress study DMEC meets at least 6-monthly and will monitor data, make recommendations and report to the independent programme steering committee.

### Patient and public involvement

A fourteen-member advisory group of people with type 1 diabetes is embedded within the D-stress programme infrastructure. They continue to advise on the intervention co-design and programme theories, the research protocol development and our knowledge translation (38).

### Setting and participants

The study is conducted in four geographically diverse UK National Health Service (NHS) specialist diabetes services at the Royal United Hospitals Bath NHS Foundation Trust, University Hospital Leicester NHS Trust, and Leeds Teaching Hospitals NHS Trust and Guy’s and St Thomas’ NHS Foundation Trust. These sites are those of clinical collaborators to represent geographical and ethnically diverse populations. Within each site, twenty adults with type 1 diabetes using a continuous glucose monitoring (CGM) device will be recruited (N=80) to the cohort study. From this cohort up to 20 will be interviewed to explore their experience of receiving EUC and/or REDUCE as part of the D-stress Pathway. Up to ten family members or friends of consenting participants and up to sixteen health professionals delivering the care pathway will also be recruited.

### Eligibility

#### Inclusion criteria

People with diabetes: Adults aged ≥18 years with a type 1 diabetes diagnosis of ≥12 months and have been using a continuous glucose monitor for ≥3 months. Family/friends: ≥18 years and their associated participant has consented to their involvement. Health professionals: Employed at one of four study sites (NHS specialist diabetes services in Leeds, Leicester, London, and Bath) and willing to undertake EUC training. REDUCE facilitators: Expertise and experience in type 1 diabetes care and agreement to be a study participant (see section REDUCE intervention arm, below).

#### Exclusion criteria

People with diabetes: Pregnant individuals and those diagnosed with type 1 diabetes within the past year will be excluded from the study as early adaptations to living with type 1 diabetes will impact on diabetes distress outcomes (39). Adults with ongoing severe mental health problems (e.g. psychosis or problematic substance use, active suicidal ideation or severe depression may be deemed ineligible by their clinical team if their current symptoms are likely to interfere with REDUCE group participation). Health care professionals who are unwilling and/or unable to take on additional workload associated with the D-stress e-learning and delivery of Enhanced Usual Care. REDUCE facilitators who do not receive Disclosure and Barring Service (DBS) approval.

### Recruitment and informed consent

Within each site, potentially eligible participants with T1D are identified by recruited diabetes health professionals trained in delivering the D-stress care pathway. The health professional introduces the study, and if interest is expressed, they seek consent to pass their contact details to the research team. Prospective participants are sent the study Participant Information Sheet (PIS) electronically via REDCAP and/or in person and offered the opportunity to discuss the study with the researcher and/or their routine diabetes clinician. If the prospective participant decides they would like to take part in the study, they have up to fourteen days to return the signed informed consent form back to the D-stress study research team. Participants can withdraw at any point without providing an explanation. For family members or friends of the PwT1D consent will first be sought from the PwT1D to invite their family member or friend to participate. The family members or friend will then be asked to provide consent for their contact details to be passed to the research team. Health care professionals willing to undertake Enhanced Usual Care training will be approached by the research team following introduction by the Site Principal Investigator. REDUCE facilitators were identified by snowball recruitment and invited to apply for this paid role. Those successfully appointed following the selection and interview process will be approached by a member of the research who will provide a study Participant Information Sheet and undertake written informed consent process.

### Interventions

#### Active Comparator Enhanced Usual Care arm (EUC)

The comparison intervention is called Enhanced Usual Care (EUC) which aims to reshape routine diabetes care so that emotional wellbeing becomes a legitimate and expected part of consultations as aligned with clinical guidelines (19, 26). EUC is delivered by diabetes healthcare professionals who have completed 4–6 hours of modular e-learning training (25). The training prepares the health care professionals to monitor and support the emotional and psychological health of people with type 1 diabetes attending their service, for example through administration of the T1-DDAS (40), discussion of the person’s responses during the consultation, and shared action planning (health professional and person with diabetes) to address identified problems. The delivery of EUC commences in routine care before any potential participants are approached. Only following a routine consultation with an EUC-trained HCP will a person be approached to participate in the study. EUC is expected to continue throughout the study for all participants irrespective of their diabetes distress status.

#### REDUCE Intervention arm

The experimental intervention is REDUCE and represents the management element of the D-stress care pathway for those with moderate to severe diabetes distress (mean item total score ≥3 on the T1-DDAS scale) (17). It is a psychological intervention aligned with clinical guideline recommendations (41). REDUCE is an online programme of six ninety-minute sessions in groups of 8-12 participants and 2-3 facilitators. It comprises six structured online group weekly sessions targeting diabetes distress in adults with type 1 diabetes. Session 1 creates a supportive environment and introduces the concept and normalisation of diabetes distress. Session 2 guides participants to identify emotional, cognitive, and social contributors to distress, distinguishing controllable from uncontrollable factors. In Session 3, participants explore personal triggers and interpretive patterns underpinning their “diabetes distress story.” Session 4 fosters compassionate, non-judgemental observation of these experiences. Session 5 supports translating insights into purposeful behavioural strategies through personalised action planning. Session 6 consolidates learning, clarifies behavioural changes, and facilitates goal-setting for continued self-management post intervention. Across all sessions, facilitators use structured discussion, reflective exercises, visual tools, worksheets, and peer learning. Each session includes routine check-ins, reflection, and structured closure. REDUCE participants select a six-week programme from a range of available start dates. Each REDUCE programme delivered will have the same participants and facilitators throughout. REDUCE is delivered by an specifically appointed core team of facilitators with professional accreditation with one of the following professional bodies: Nursing and Midwifery Council, Health and Care Professions Council, British Association for Counselling and Psychotherapy, or UK Council for Psychotherapy, and/or, have a professional or personal understanding of all of the following: type 1 diabetes, NHS diabetes care and guidelines, diabetes distress, mental and emotional health, professional experience of in-person and/or online group facilitation, especially in managing diverse needs within a group and the expression of strong emotions; and a willingness to be a research participant. Selected facilitators complete REDUCE training and demonstrate competency over 16 hours across five training sessions.

## STUDY PROCEDURES

Each site will identify between 1-3 diabetes clinics (clinician sessions) within which the D-stress care EUC pathway is delivered. Limiting clinical sessions to 3 per week enables the same sites to participate in the main study through up to 7 additional weekly clinics. This restriction aims to reduce the impact of contamination between those HCPs participating in the feasibility trial and those who will participate in the main trial. Once the participating HCPs complete the D-stress e-learning, they begin delivering EUC within the nominated clinic. Figure 1. illustrates the study design.

### Cohort procedures

Once informed consent is completed, monthly data collection of diabetes distress and interstitial glucose Time in Range (TIR) commences. This continues until the site has achieved it target cohort sample size following which baseline data collection for all participants is undertaken. Monthly data collection continues for the duration of the study. All participants repeat baseline measures at the completion of the trial period.

### Trial procedures

The first two sites to recruit to cohort target of 20 PwT1D per site will be manually randomised to receive the REDUCE experimental intervention first or second. We note that the randomisation does not serve a useful inferential purpose given the sample size but is used to test the procedures of the full study. In line with the proposed cluster trial design, each “batch” includes a baseline and endline period with respect to the REDUCE intervention such that all eligible patients receive the intervention by the end of the study. The remaining two sites will follow the same procedure two months later. Participants with baseline elevated diabetes distress will be offered REDUCE programme participation as per their cluster sequence. REDUCE programs may therefore comprise participants from any of the participating sites across the later trial periods. Participants with baseline sub-threshold diabetes distress continue to receive EUC and monthly data collection. If their diabetes distress elevates subsequently, they will be offered the REDUCE programme for participant safety reasons, but their data will not be included in the REDUCE trial analysis (see Supplemental material 1).

### Realist process evaluation procedures

Participants with diabetes and consenting family/friends and D-stress delivery health care professionals will be purposively sampled for interview or observations of their clinic consultation. D-stress delivery health professionals and facilitators will be interviewed, and their care delivery observed as soon as possible following commencement of the delivery of EUC or REDUCE. Interviews and observations will continue throughout the whole study period including participants with both elevated and sub-threshold diabetes distress in both the cohort and the trial (see Fig 1).

### Outcome and assessments

The data collection methods and schedule for study assessments and outcomes are presented in Table 1.

**Table 1:** Summary of assessments for cohort and trial participants. ✔ indicates the assessment is collected at specified timepoint.

| Outcome | Measure/ Instrument | When collected | How collected | Cohort assessment | Trial assessment |
| --- | --- | --- | --- | --- | --- |
| **Glycaemic level** | Interstitial glucose Time in Range (TIR), Time Below Range (TBR), Time Above Range (TAR), and glycaemic variability | Every 30 days | CGM sensor permission provided to research team | ✓ | ✓ |
| **Diabetes Distress** | T1-DDAS 8 item Core scale | Every 30 days | Self-report | ✓ | ✓ |
| **Diabetes Distress** | T1-DDAS 30 item Core and Source combined scale | Baseline and end-of-study follow-up | Self-report | ✓ | ✓ |
| **Demographics** | Bespoke D-stress study Demographic Questionnaire | Baseline | Self-report | ✓ | ✓ |
| **Social Support** | Berlin Social Support Scale (Schulz & Schwarzer, 2003) | Baseline and end-of-study follow-up | Self-report | ✓ | ✓ |
| **Quality of Life** | EQ-5D-5L Quality of Life measure (Kreimeier, Astrom et al. 2019) | Baseline and end-of-study follow-up | Self-report | ✓ | ✓ |
| **Loneliness** | UCLA Loneliness Scale (continuous) (Russell, 1996) | Baseline and end-of-study follow-up | Self-report | ✓ | ✓ |
| **Acceptability, Appropriateness, Feasibility** | AIM, IAM & FIM (Weiner, Bryan et al., 2017) | Baseline and End-of-study follow-up | Self-report | ✓ | ✓ |
| **Health & Social Equalities** | Bespoke D-stress study questionnaire categorical questions (e.g., disability, food/fuel security) | Baseline and end-of-study follow-up | Self-report | ✓ | ✓ |
| **Limited Healthcare Use** | Number and dates of hospital admissions/emergency events | End-of-study follow-up | Electronic health record | ✓ | ✓ |

#### Denographic and medical history assessment

To characterise the population participating in the study, socio-demographic and clinical information will be collected from all participants enrolled in the cohort, including those with both sub-threshold and elevated diabetes distress at baseline. These data will include age, gender, ethnicity, education, occupational status, co-habiting status, and lifestyle characteristics, such as smoking. Clinical variables will include diabetes duration and diabetes management approach (e.g. pump, hybrid closed loop, or multiple daily injections). These variables will be used to describe the study population, support subgroup and moderator analyses, and inform purposive sampling for the process evaluation.

#### Cohort assessments

Cohort assessments are collected at participant recruitment to the cohort and monthly until the final follow up in month 7 (Table 1).

##### Diabetes Distress assessment

Participants diabetes distress score is assessed using the T1-Diabetes Distress Assessment System (T1-DDAS) Core scale (17). The DDAS Core scale is an 8-item assessment of overall diabetes distress. A core score of ˂3 on each item we determine sub-threshold and ≥3 as elevated distress.

##### Interstitial glucose Continuous Glucose monitoring

We will obtain monthly routine glucose metrics from participants usual CGM systems including CGM usage (30 days), time in range (TIR; glucose between 3.9-10.0 mmol/L), time above range (TAR), (level 1 and 2), time below range (TBR) (level 1 and 2); and coefficient of variation (CV). Monthly scan data will be accessed from the participant’s CGM account at the end point by the research team only and entered into REDCap.

#### Trial assessments

Trial assessments are collected at the baseline point of month 3 for cluster batch 1 and month 5 for cluster batch 2. The final follow up for both batches is month 7 (see Table 1).

Diabetes distress is a proposed co-primary outcome for the main trial. The T1-Diabetes Distress Assessment System (DDAS) scale determines eligibility for the REDUCE programme trial. The proportion of interstitial glucose time in range is a proposed co-primary outcome for the main trial collected as part of the cohort study data collection.

#### Other trial outcomes are

- Proportion of interstitial glucose time above range (TAR) and time below range (TBR) data will be assessed from their routinely used continuous glucose monitoring devices.
- Level of social and cognitive support in everyday life assessed by Berlin Social Support Scale (BSSS) tool (see Table 1 for secondary outcome measurement tools) (42).
- Perceived health related quality of life across five domains: mobility, self-care, usual activities, pain / discomfort, and anxiety / depression assessed using the EQ-5D-5L (12).
- Subjective experiences of loneliness and self-perceptions of social connectedness measured using the UCLA Loneliness Scale (43).
- Participants subjective experience and perception of acceptability, suitability, feasibility and appropriateness of study intervention (44).
- Asset of food and fuel insecurity, and available social support and disability assessed by a D-stress study specific questionnaire (see Supplemental material 2)
- Number and dates of hospital admissions and emergency events, such as events related to severe hypoglycaemia for participant. These data will be extracted directly from participants’ electronic medical records to ensure accuracy and completeness.

#### Feasibility assessments

- Proportion of people recruited and retained to the cohort, and to the trial, from those eligible and approached through calculation of those who consent over the number of Participant Information Sheets (PIS) sent to eligible participants who subsequently consented.
- Number of participants who drop out of the research and the REDUCE programme including reasons to be determined by quantitative documentation of dropout rates and reasons and realist process evaluation in-depth interviews.
- Feasibility and acceptability of data collection determined by analysis of questionnaires and CGM scans, and missing data.
- Quality of outcome data assessed through quantitative evaluation of missing data patterns.
- Retention rates across study site/ milestones.
- Diversity of recruited participants against nine protected characteristics determined through analysis of study questionnaires completed at baseline and end of study data collection points.
- Determination of whether progression to main trial is possible, determined through mixed methods analysis related to prespecified Progression Criteria (see Table 2).

### Data collection

All patient reported outcome data for the cohort and trial are collected electronically via the King’s College London licensed Research Electronic Data Capture (REDCap) software platform. Participants receive by email or text a link to a single monthly survey and additional survey links at baseline and follow up to complete assessments. Participants’ routine CGM sensor, linked to their sensor providers portal, is accessed by the research team with the participant’s consent to download glucose data.

**Table 2:** Progression Criteria.

|  | Feasible | Feasible with<br>Protocol modification | Not feasible |
| --- | --- | --- | --- |
| Identify, recruit, and train minimum 2 HCPs (per site) | ≥12 participants (75%) | 8-11 participants | ≤8 participants |
| Train HCP to deliver EUC to recruit 20 PwT1D participants per site (n=80) | ≥10 (50%) participants will have DD ≥3 and eligible for REDUCE | ≤20 participants | ≤ 7 participants |
| Retention of participants with sub threshold and elevated DD | ≥60 participants (75%) | 40-59 participants across 4 sites (50%) | ≤ 40 (≤ 50%) participants |
| CGM device data scanned/downloaded | ≥60 participants CGM data downloaded (75%) | 40-59 participants CGM data available (50%) | ≤40 participants CGM data downloaded (≤50%) |
| Completeness of co-primary outcome data (DD score) | ≥60 participants (75%) | 40-59 participants (≥50%) | ≤ 40 participants (≤ 50%) |
| Completeness of secondary outcome data: completion of REDUCE programme | 8-10 participants attend four of six sessions | 5-7 participants complete 4 out of 6 sessions | ≤ 4 participants complete 4 out of 6 sessions |
| PwT1D with DD ≤ 3 consenting to realist interviews/observations (per site) | ≥5 participants | 3-5 participants | ≤3 participants |
| PwT1D with DD ≥3 consenting to realist interviews/observations (per site) | ≥5 participants | 3-5 participants | ≤3 participants |
| HCPs consenting to interviews/observations (per site) | 3-4 participants | 2-3 participants | ≤ 2 participants |

**Legend:** Progression criteria will be assessed by a traffic-light approach. Green indicates proceed to main trial; Amber indicates protocol modifications required; Red indicates criteria have not been met and substantial revisions required.

#### Realist process evaluation data collection

Qualitative data collection continues between months 1 and 7 month (25, 45). The outcomes of interest are not whether the EUC and REDUCE interventions ‘worked’ but how, why, for whom, in what contexts and under what circumstances outcomes were produced. Realist process evaluation follows RAMESES quality and publication standards to conduct semi-structured one-to-one interviews, and non-participant observation of clinical encounters between patient and health care professionals and online REDUCE intervention programmes (46, 47). These will be undertaken at various timepoints across the cohort and trial participant population. Observations and interviews will be audio or video recorded in-person, on Microsoft Teams, or by telephone and arranged at a time convenient to participants.

#### Data management

To protect confidentiality before, during and after the trial, all personal information collected about potential and enrolled participants will be stored in a secure password protected Master File stored on the King’s College London server for a maximum of ten years. All study participant names are pseudonymised.

#### Data sharing

Individual de-identified participant data and any other materials will be accessible on REDCap by the research team who will have specific user permissions in place.

### Sample size

To evaluate the feasibility of the D-stress study pathway we aim to recruit 80 participants, (20 per clinic) and hence expect to invite 240. This is based on estimating the confidence interval around the proportion of participants that we expect will be retained in the trial. We assume 25% of participants might drop out, with 75% being retained in the study. 80 patients will allow to estimate the proportion of patients retained in the study with a precision of approximately +/- 10 percentage points as a 95% credible interval assuming a mean retention of 75%. If the retention is as high as 95% then the 95% credible interval will be about +/-5 percentage points

A consent rate of 30%, a key feasibility study outcome, is anticipated to be sufficient because psychological and emotional health care pathways are amongst the greatest in terms of unmet needs voiced by people with type 1 diabetes (5). Of the 80 participants we recruit; we expect approximately 40 participants (50%), approximately ten from each NHS site, will have elevated DD and will be invited to participate in REDUCE (17, 28). The sample of 40 participants with elevated DD will be sufficient to estimate recruitment and retention rates and data quality, which are primary research questions. The realist evaluation team will purposively sample up to 20 participants for interviews and up to 20 observations of clinical consultations and REDUCE sessions. Participants with sub-threshold or elevated diabetes distress scores will be selected to reflect a range of socio-cultural demographic and clinical characteristics. The proposed sample size was selected to provide sufficient data to assess feasibility outcomes, including recruitment, retention, intervention delivery and data completeness, rather than to detect statistically significant treatment effects

#### Randomisation

Manual randomisation occurs at the cluster (site) level. The first two clusters (sites) to have recruited their target participants according to the available time frame will be randomised to receive the REDUCE programme first or second, one month apart. These two clusters become Pair 1. Clusters 3 and 4 form Pair 2 and will follow the same randomisation procedure when ready (Fig 1). The senior trial statistician will generate random allocation sequence.

#### Blinding

This is an unblinded study. Due to the need for the research and clinical teams to interact with participants about their involvement in REDUCE and its associated data collection, it is not possible to mask participants, the site health care teams or the researchers.

#### Harms

The D-stress study will follow the Safety Protocol (Supplemental material 1) written for participants and PPIE study members. The King’s Health Partners Serious Adverse Events template will be used across all study sites.

## ANALYSIS

The feasibility of implementing the D-stress main cluster randomised trial will be reviewed after the completion of the feasibility study and assessed according to the progression criteria outlined in Table 2.

### Statistical analysis

The purpose of the feasibility study is to test feasibility of recruitment, retention and intervention delivery and provide estimates (means/ median and estimates of variance) of key planned primary/ secondary outcomes that will be used to support the design of the main study evaluation. The proposed analyses are therefore descriptive and exploratory analyses, without formal statistical testing. Continuous analysis (e.g. of age, distress score) will be summarised by mean, standard deviation, and 95% confidence interval, and binary outcomes (recruitment proportion and elevated DD prevalence). Patient-level outcomes, including socio-demographic, medical history, and main trial primary outcomes, will be summarised at the cluster level and overall. The main trial outcomes will also be summarised by time period. Cluster-level outcomes, including recruitment and dropout rates, will be summarised by time period and overall. Tables will be produced for all summaries with graphical presentation of the results where appropriate.

### Realist process evaluation analysis

Analysis will focus on evaluating how the programme theories (45) can be used to (a) explain why and how EUC and REDUCE study interventions might or might not work, and (b) refine the context, mechanism, outcome configurations (CMOs) of each intervention in preparation for evaluating these in the main randomised clinical cluster trial. Interview and observation data will be transcribed and analysed using framework analysis (48) to identify themes within the data and to facilitate comparison between clusters and participants. An analytical framework developed from the programme theories identified in the rapid realist reviews will allow qualitative data to be analysed systematically while also accommodating the identification of new themes and refinements to the initial programme theories. Attention will be given to identify new mechanisms found in the data, not previously identified in the realist reviews, which are considered to explain how the interventions work or not. The framework approach can be used to manage qualitative data and undertake analysis systematically, ensuring the analyser maintains an explicit audit trail (49). This enhances the rigour of the analytical process and encourages a greater confidence in the credibility of the findings (45). Data analysis will be undertaken throughout the trial period.

### Participant safety protocol

A study specific safety protocol has been designed to address risks that participation in the feasibility study might trigger in relation to emotional discomfort and distress, which raise safety concerns (see Supplemental material 1).

## Supporting information

Supplemental material, Safety Protocol

Supplemental material, Health and social equalities questionnaire

## Data Availability

All data produced in the present study are available upon reasonable request to the authors.

https://www.kcl.ac.uk/research/d-stress-study

## Author declarations

The following authors have conflicts of interest SW, PC, JS, Rha, and RIGH. All other authors have no conflicts of interest to declare.

## Role and contact information of trial sponsor

King’s College London and Guy’s and St Thomas’ NHS Foundation Trust are trial co-sponsors. Correspondence to.

## Author Contributions (CRediT)

**Conceptualization:** SA., RIGH., SH., PC., RHa., FF., MDC., JHu., LF., KH., JHa., IPN, VS., RA. EL. **Data Curation:** CFT, RHa, MP, JM, SS. **Formal Analysis:** CFT, SA, SW, SH, PC, RHa, MP, SS, MDC, VS. **Funding Acquisition**: RIGH., SH., PC., RHa., FF., MDC., JHu., LF., IPN., VS., RA., G H. **Investigation:** CFT, SA., SW., SH., PC., RHa., MP., SS., MDC., LF., JHa, RW., VS. **Methodology:** RIGH., SW., SH., PC., RHa., FF., MDC., JHu, LF., KH, HJ., RW., VS. **Project Administration:** CFT., MP., JM., SS., RW., GH. **Resources:** CFT., MP., JM., SS., JHa. EL **Software**: SW., HJ. **Supervision:** RHa., LF., **RA., GH. Validation:** HJ., MS., GH. **Writing – original draft:** CFT, SA. **Writing – review & editing:** CFT., SA., RIGH., SW., SH., PC., RA., RHa., MP., JM., SS., FF., MDC., JHu., LF., KH., MS., HJ., J.S., Jha., RW., IPN., VS., RA., GH.

## Acknowledgments

With thanks to our D-stress study patient forum, and Expert Reference Group, whose insight and expertise has been invaluable in the design of the study. We are most grateful to those who agreed to participate in the trial. Your continued support has made this study possible.

