## Supplemental material, Safety Protocol for "Optimising the prevention, detection and management of diabetes distress in adults with type 1 diabetes: Feasibility study protocol (D-stress feasibility study)"

### Supplemental material 1

#### Participant Safety Protocol

The safety of potential and actual participants is the greatest ethical concern and there is a risk that participation in the feasibility study might trigger emotional discomfort, distress, and raise safety concerns. In addition to the safety of study participants the safe and effective management of patient and public involvement (PPIE) members of the D-stress study research team is also of greatest ethical concern. Should the PPIE Co-investigator on the D-stress programme and a Senior Public Involvement Manager be aware of mental health concerns of PPIE members, the Feasibility Safety Protocol for study participants will be followed.

Our feasibility Safety Protocol will address the safety needs of participants and across the study. It follows the best practice, professional guidelines, and local NHS policies for monitoring mental state and risk throughout the participants' involvement in the study and will be facilitated by close liaison with research and clinical teams. The safety of the feasibility study, which involves Enhanced Usual Care (EUC) and trial which involves EUC and the REDUCE intervention, will be monitored closely during clinical consultations and online REDUCE sessions and through regular contact with the participant's clinical team.

The following comprises our safety protocol for all participants in the study: those participants receiving Enhanced Usual Care, those participating in the feasibility trial randomised to receive the comparator and experimental intervention and those that withdraw. It addresses lines of responsibility and accountability, definitions relating to safety, escalation and safeguarding procedures in the event of notable clinical deterioration with or without an escalation in risk.

##### 1.0 Lines of responsibility and accountability

1.1. Throughout a person's participation in the D-stress feasibility study, their routine diabetes clinician holds responsibility and accountability for taking action in relation to the participant's mental health and wellbeing, and this includes their safety.

1.2. For those participants recruited to the feasibility study from the point at which the participant consents to join the study until their final data collection point, responsibility for monitoring their diabetes distress and escalating care as a consequence of observing elevating diabetes distress, lies with the research team. This includes observations made during data collection, data analysis, or REDUCE programme participation.

1.3 Participant safety will be a standing agenda item on every fortnightly research team's Work Package group meeting.

#### 2.0 Definitions related of safety

2.1. Diabetes Distress – the emotional and psychological burden associated with living with the disease. Symptom and degree of diabetes related distress and severity is assessed using the T1-DDAS Core, Source or Combined scale. The T1-DDAS is a self-report screening questionnaire completed by the participant at the beginning of the feasibility study and thereafter each month of the study until discharge. Any existing or emergent safeguarding and /or vulnerable adult concern will be assessed and monitored at each clinical consultation (whether that takes place in person or remotely) with proportionate action taken in accordance with legislative reporting requirements. Elevated diabetes distress is a score of ≥3 on this scale. We define concerning DD indicating possible co-morbidity with other mental health conditions such as depression as a score of ≥4.

2.2. The risk of Self-Harm Adverse Event (see Section 10) is defined as when the research and/or clinical team has been unable to make contact on three separate occasions over three days with a participant scoring ≥4 on the DDAS within the previous month, following engagement with any D-stress research procedures.

#### 3.0 Safety net procedures for all participants

3.1 All participants will receive a Participant Information Sheet. This will list the contact details of the clinical trial manager as well as independent services to call 24/7 if they feel they need to talk with someone about their mental health throughout their study participation.

3.2. For participants enrolled in REDUCE or data collection the facilitator or researcher will record any concerns in the D-stress study participant safety log, contact the trial manager or their delegate and escalate according to 2.2.

3.3. The trial manager will report to the participant safety standing item at every two-weekly team meeting.

#### 4.0 Care escalation procedures in the event of a risk of self-harm

4.1. When a risk of a Self-Harm Adverse Event is identified (see 2.2) the trial manager or their delegate will contact the participant using their usual preferred contact method to check in with them on their wellbeing and to suggest they may wish to speak to their GP for support. 4.2. If the research team assessor is unable to make contact or do not hear back from the participant within 24 hours, they will send a follow-up email.

4.3 If research team assessor still receives no response, they will send a third communication saying they have emailed the participant twice over the past two days and if they do not hear from them within 24 hours, they will get in touch with their routine diabetes team to let them know that they are concerned about them.

4.4. If this final communication does not elicit a response, the research team assessor will escalate the reporting and contact the participants routine diabetes care team to voice our concerns. This will be reported to the study's senior investigator (Prof Sturt) and be registered as an adverse event.

#### 5.0 Care escalation procedures in the case of adverse event

5.1. If at point of referral or during the course of treatment, research and /or clinical teams become concerned about the welfare of any participant or immediate family member, or PPIE member, they will escalate their concerns through D-stress study's standardised risk assessment, escalation, management and safeguarding policies and procedures. Likewise, if a safeguarding and vulnerable adults concern is identified this will be escalated, acted on and reported to the relevant statutory body – safeguarding team.

#### 6.0 Participants that withdraw

6.1. Participants that join the study are free to withdraw at any point. In this circumstance the monitoring of the person's mental health and wellbeing revert to the participant's GP and/or routine diabetes care team.
6.2. Once the research team is aware of the participant's withdrawal, they will contact the participant to thank them for taking part in the study and will provide them with tailored individual signposting to mental health services, where appropriate.
