## Supplemental material, Health and social equalities questionnaire for "Optimising the prevention, detection and management of diabetes distress in adults with type 1 diabetes: Feasibility study protocol (D-stress feasibility study)"

### Supplemental material 2

#### Health and social equalities questionnaire

1. **Disability** (tick all that apply)
   a) Autism/autistic spectrum
   b) Developmental (e.g. affecting motor, cognitive, social, language)
   c) Hearing (e.g. D/deaf, partial loss)
   d) Learning disability
   e) Learning difference (e.g. dyslexia, dyspraxia, ADHD)
   f) Long-term physical illness/ health condition apart from diabetes (e.g. cancer)
   g) Mental health difference/condition (e.g. depression, schizophrenia)
   h) Physical difference (e.g. condition limiting basic physical activities)
   i) Sight (e.g. blindness, partial sight loss)
   j) Speech and language
   k) Visible difference with a disabling and/or discriminatory impact
   l) Something else (please specify)
   m) Prefer not to say

2. **Co-habiting**
   a) Do you currently live with others?
      a. Yes / No
   b) How many people typically live in your household?
      a. Open-ended text field or drop-down menu
   c) Who do you live with? (Check all that apply)
      - I live on my own
      - Spouse/partner
      - Children
      - Parents
      - Siblings
      - Other relatives
      - Friends/family friends
      - Roommates
      - Others (specify)

3. **Housing/Residency**

What best describes your current living arrangement? [Select one]
   - Own house/apartment (with or without mortgage)
   - Part own house/apartment (shared ownership)
   - Rent apartment/bedsit/house

- Live rent-free with parents/family/friends
- Temporary accommodation (hotel/bed-and-breakfast/sofa-surfing/shelter)
- Homeless
- Other (please specify)

**4. Fuel poverty**

a) Do you feel stressed or anxious about your ability to heat your home to the temperature you would like?
b) Have you experienced any of the following in the past month due to fuel costs?
   a. Difficulty concentrating
   b. Feeling irritable or short-tempered
   c. Trouble sleeping
   d. Loss of appetite
   e. Feeling overwhelmed or unable to cope
c) How satisfied are you with the warmth in your home? Scale from 1 to 5
d) Have you had to use any of the following coping strategies to stay warm?
   a. Wearing extra layers of clothing
   b. Using blankets or sleeping bags
   c. Taping over vents
   d. Co-sleeping with children

**5. Food insecurity**

In the last 12 months, can you tell me if these statements were true for you?

1 “We worried whether our food would run out before we got money to buy more.”
Often true Sometimes true Never true

2 “The food that we bought just didn’t last, and we didn’t have money to get more.”
Often true Sometimes true Never true

3 “We couldn’t afford to eat balanced meals.”
Often true Sometimes true Never true

We could ask further questions as suggested by ENUF, but at most we could add this question:

Stage 2 Adult/Households questions (if one or more Stage 1 Adult/Household questions affirmed)

In the last 12 months…

4a Did (you/you or other adults in your household) ever cut the size of your meals or skip meals because there wasn’t enough money for food? Yes No

4b If yes: How often did this happen—almost every month, some months but not every month, or in only 1 or 2 months?

Almost every month Some months but not every month Only 1 or 2 months

6. **Financial Stress**
    a) On a typical day, how concerned are you about your finances? (Scale: 1-5, where 1 is 'not concerned at all' and 5 is 'extremely concerned')
    b) Have you experienced any sleep disturbances or anxiety related to financial matters in the past month?
    c) How often do you experience feelings of financial overwhelm or burnout? (Scale: 1-5, where 1 is 'never' and 5 is 'daily')
